# Validation of Diabetes Prediction Scores: Does adding a high risk for depression increase the area under the curve?

**DOI:** 10.1101/2023.11.30.23299228

**Authors:** MA Salinero-Fort, J Mostaza, C Lahoz, J Cárdenas-Valladolid, V Iriarte-Campo, E Estirado-de Cabo, F García-Iglesias, T González-Alegre, B Fernández-Puntero, V Cornejo-del Río, V Sánchez-Arroyo, C Sabín-Rodriguez, S López-López, P Gómez-Campelo, B Taulero-Escalera, F Rodriguez-Artalejo, FJ San Andrés-Rebollo, C de Burgos-Lunar, SPREDIA-2 Group

**Affiliations:** Biosanitary Research and Innovation Foundation of Primary Care (FIIBAP); Frailty, multimorbidity patterns and mortality in the elderly population residing in the community - Hospital La Paz Institute for Health Research IdiPAZ, Madrid, Spain; Network for Research on Chronicity, Primary Care, Health Promotion (RICAPPS) Madrid, Spain; Lipid and Vascular Risk Unit, Department of Internal Medicine, Hospital Carlos III, Madrid, Spain; Department of Preventive Medicine, San Carlos Clinical University Hospital, Madrid, Spain; Miraflores, Primary Health Centre, Madrid, Spain; Alfonso X El Sabio University, Madrid, Spain; Hospital Carlos III, Madrid, Spain; Hospital La Paz Institute for Health Research (IdIPAZ), Madrid, Spain; Department of Preventive Medicine and Public Health, Universidad Autónoma de Madrid. CIBERESP (CIBER of Epidemiology and Public Health), and IMDEA-Food Institute, CEI UAM+CSIC, Madrid, Spain; Las Calesas Health Centre, Madrid Spain; Hospital Clínico de San Carlos, Madrid, Spain

**Keywords:** Diabetes Mellitus, Risk Factors, Surveys and Questionnaires, ROC Curve

## Abstract

**Background:** Diabetes risk scores include age, waist circumference, body mass index, hypertension, use of blood pressure medication, and metabolic and lifestyle variables. Although patients with major depressive disorder have a higher risk of diabetes, none of the diabetes risk scores includes high risk of depression as an additional item.

**Aim:** To validate three diabetes risk scores (FINDRISC, DESIR, ADA) in the Spanish population aged >45 years with the aim of predicting diabetes and to test the value of adding high risk of depression, defined as a PHQ-9 questionnaire score ≥10, to the risk score with the best discriminative performance.

**Methods:** Prospective population-based cohort study in Madrid (Spain). FINDRISC, DESIR, ADA, PHQ-9, and OGTT values were measured at baseline. Participants with OGTT <200 mg/dl (n= 1,242) were followed up for a median of 7.3 years using their general practitioner’s electronic health record (EHR) and telephone contact. Incident diabetes was identified as treatment for diabetes, fasting plasma glucose ≥126 mg/dl, a new diagnosis in the EHR, or self-reported diagnosis. At the end of the study, the performance of diabetes risk scores, including a modified original FINDRISC score with a new variable for high risk of depression (FINDRISC-MOOD), was assessed.

**Results:** During follow-up, 104 (8.4%; 95% CI, 6.8-9.9) participants developed diabetes, and 185 had a PHQ-9 score ≥10. The AUROC values were 0.70 (95% CI, 0.67-0.72) for FINDRISC-MOOD and 0.68 (95% CI, 0.65-0.71) for the original FINDRISC. The AUROC for DESIR and ADA were 0.66 (95% CI, 0.63-0.68) and 0.66 (95% CI, 0.63-0.69), respectively. There were no significant differences in the AUROC between FINDRISC-MOOD and the remaining scores.

**Conclusion:** FINDRISC-MOOD performed slightly better than the other risk scores, although the differences were not significant. FINDRISC-MOOD could be used to identify the risk of future diabetes.

## Introduction

According to the International Diabetes Federation (Diabetes Atlas, 2021), diabetes mellitus (DM) affects approximately 537 million adults (20-79 years) worldwide, of whom more than 61 million live in Europe. The prevalence of diabetes is increasing in all age groups, mainly owing to the increasing frequency of overweight and obesity (1), unhealthy lifestyle and diet (2), physical inactivity (3), and psychosocial factors such as depressive disorders (4).

DM is associated with a high risk of cardiovascular events (5), chronic kidney disease (6), and all-cause and cardiovascular mortality (7). Affected patients have often developed subclinical atherosclerosis when they are diagnosed with DM, thus increasing the risk of cardiovascular events (8).

Diabetes risk scores (FINDRISC (9), DESIR (10), ADA (11)) help to identify individuals who require laboratory measurements (eg, fasting plasma glucose, HbA1c, and the oral glucose tolerance test [OGTT] values) to be taken in order to enable populations to be stratified by prognosis. Such an approach could facilitate the implementation of interventions aimed at halting or delaying the onset of DM and preventing cardiovascular complications.

To our knowledge, no diabetes risk scores include psychosocial factors, although it is well known that patients with major depressive disorder have a higher risk of type 2 DM (T2DM), as highlighted in a recent meta-analysis (12). Therefore, we postulate that adding the high risk of depression, defined as a Patient Health Questionnaire-9 (PHQ-9) score ≥10, to the diabetes risk score with the best area under the receiver operating characteristic curve (AUROC) in our population could better identify the risk of T2DM.

The present study aims to validate three diabetes risk scores (FINDRISC, DESIR, ADA) in the Spanish population to predict the incidence of T2DM after a long follow-up period (median 7.3 years) and to test the value of adding high risk of depression to the diabetes risk score with the best discriminative performance.

## Material and Methods

### Design

This study was conducted as part of a broader project funded by the Spanish Instituto de Salud Calos III (PI 1500259). It included the Screening PRE-diabetes and type 2 DIAbetes (SPREDIA-2) study, which has been described in detail elsewhere (13). SPREDIA-2 is a population-based prospective cohort study, in which baseline visits were scheduled from July 2010 to March 2014.

### Population

#### Baseline visit

The study population comprised a random sample of 2,553 subjects living in the north of the city of Madrid (Spain) in an area served by 10 primary health care centers. Of these, 1,592 (62.4%) agreed to participate, and 1,426 had not been previously diagnosed with DM.

Recruitment was divided into three stages:

1. Potential participants were sent a letter signed by their general practitioner explaining the aims of the study and inviting them to participate.
2. Subjects were contacted by telephone to clarify any doubts and, if interested, were given an appointment to be assessed.
3. The patient attended the assessment at the Carlos III Hospital outpatient clinic after an overnight fast.

A fasting blood sample was taken upon arrival at the outpatient clinic to determine levels of glucose, creatinine, uric acid, HbA1c, serum insulin, lipids, and lipoproteins. Immediately after blood sampling, all subjects not previously diagnosed with DM underwent an OGTT with 75 g of anhydrous glucose in a total fluid volume of 300 ml. A second blood sample was taken 2 hours later. The measurement questionnaires were as follows: diabetes risk scores (FINDRISC, DESIR, and ADA); the PHQ-9 (14); the 14-item Questionnaire to assess adherence to the Mediterranean diet (PREDIMED) (15); and the 12-item Short-Form Health Survey (16). Anthropometric measurements and a full clinical history were taken. Alcohol consumption was measured as the number of units of alcohol per week.

### Follow-up

Participants were followed up for a median of 7.3 years between the baseline visit and 31 December 2019 using their general practitioners’ electronic health records (EHRs). The EHRs had previously been validated (17) and used in epidemiological studies (18). The participants were also contacted by telephone during the last year of follow-up to ascertain whether they were alive and, if so, to record health status, including the incidence of T2DM or cardiovascular events. The interview was conducted by a researcher trained in obtaining medical data by telephone. The study flow chart is shown in Figure 1.

**Figure 1.**
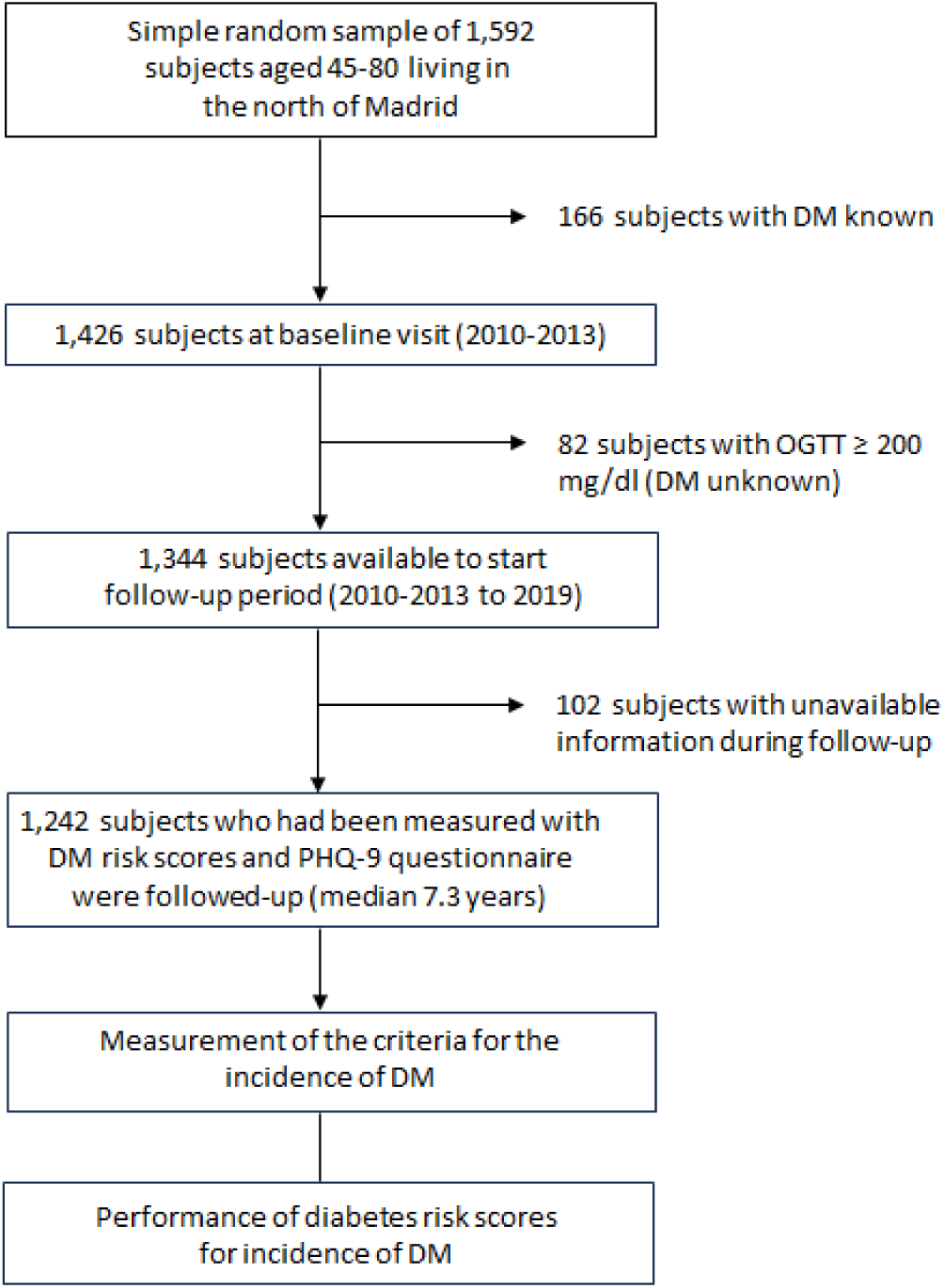
Study Flow-Chart.

### Measurement tools and definitions of criteria

FINDRISC risk score: The Finnish Diabetes Risk Score is one of the most widely used (9). It includes 8 variables (anthropometric and lifestyle), namely, age, body mass index (BMI), waist circumference, family history of diabetes, use of blood pressure medication, history of high blood glucose levels, daily physical activity, and daily intake of vegetables, fruit, and berries. FINDRISC assesses the likelihood of developing T2DM over the next 10 years. The score ranges from 0 to 26 points, and the usual cut-off is 15. A risk score of 0–14 points indicates a low-moderate risk of diabetes (1–17% risk of diabetes over 10 years), and 15–20 points indicates a high risk of diabetes (33% risk of diabetes over 10 years). A score of 20–26 points indicates a very high risk of diabetes (50% risk of diabetes over 10 years) (19).

DESIR risk score: DESIR was designed by Balkau et al (10) in the French population. The component variables differ by sex. In women, the variables include waist circumference (cm), family history of diabetes, and arterial hypertension, while for men, they include waist circumference (cm), current smoking status and arterial hypertension. The waist circumference categories differ by sex (for women, <70, 70-79, 80-89, ≥90; for men, <80, 80-89, 90-99, ≥100). The score ranges from 0 to 5 points (a higher score means a higher risk).

ADA risk score: The American Diabetes Association risk score (11) was developed based on the US population older than 20 years without DM to identify individuals at high risk for DM or prediabetes. It includes the following variables: age, sex, race, weight, height, family history of DM, history of gestational DM, history of arterial hypertension, and physical activity. The total score ranges from 0 to 11. A score of five or higher indicates a high risk of DM (11).

PHQ-9: This validated and reliable scale has been used in many research studies (20). It showed high internal consistency at both time points (Cronbach’s alpha = 0.71 Pre; 0.77 Post) and a significant test-retest correlation (r _paired_ = 0.50). The question put is “Over the last 2 weeks, how often have you been bothered by any of the following problems?”, with four response options: (0) Not at all; (1) Several days; (2) More than half the days; and (3) Nearly every day. The total PHQ-9 score is assessed by adding together the scores for all nine items. Higher scores on this measure indicate greater depression. Scores are categorized into five levels of severity: minimal = 0 to 4; mild = 5 to 9; moderate = 10 to 14; moderately severe = 15 to 19; and severe = 20-27. The optimal cut-off for major depression disorder is 10 (21). It takes approximately 2-5 minutes to administer. Additionally, it can be self-administered (22).

FINDRISC-MOOD risk score: This is an adaptation of the original FINDRISC risk score in which five new points are added if participants scored a positive PHQ-9 for depression (≥10 points). In contrast, no points are added if the PHQ-9 is negative. We decided that five points was appropriate based on the value of the beta coefficient obtained in the PHQ-9 after adjustment for the FINDRISC score in the logistic multivariate analysis for the prediction of diabetes (beta coefficient=0.632). This coefficient was multiplied by nine, which was the smallest common multiplication factor possible to obtain a sensitive score.

### Diagnostic criteria

Incidence of DM (Gold standard): Incident cases of diabetes were identified by treatment for diabetes, fasting plasma glucose ≥126 mg/dl, new diagnosis in the EHR (T90 code from the International Classification of Primary Care, Second Edition), or self-reported diagnosis in the telephone interview.

Metabolic syndrome: Metabolic syndrome was defined according to ATPIII diagnostic criteria (23).

### Statistical methods

The statistical analyses were conducted using IBM SPSS Statistics for Windows, Version 26.0 (IBM Corp, Armonk, New York, USA) and MedCalc for Windows, version 15.8 (MedCalc Software, Ostend, Belgium). The sociodemographic and clinical characteristics of the study population at baseline are presented as frequencies and percentages for categorical variables and as means and standard deviations (SDs) for continuous variables. Between-group comparisons were performed using a chi-square or Fisher’s exact test for categorical variables and a *t* test or Kruskal-Wallis test for continuous variables.

To calculate the sample size, the following assumptions were accepted: an α error of 0.05, a precision rate of 9% in a bilateral contrast, for an estimated specificity rate of 80% and an estimated incidence of DM of 6%; the total sample size required was 1217 participants.

The performance of the diabetes risk scores was assessed using the following indicators: sensitivity, specificity, positive and negative predictive values, Youden index defined as [sensitivity + specificity –1], and positive and negative likelihood ratios. All scores were calculated using incident T2DM as the gold standard. Statistical significance was set at p < 0.05 for a two-tailed test.

The discriminative accuracy of the different risk scores was assessed and expressed as the AUROC and corresponding 95% CIs. AUROCs were compared between scores using MedCalc software.

The cut-off points for the risk scores to identify incidence of T2DM were determined by the point with the shortest distance to the upper left corner of the ROC curve, as calculated using the Youden index.

## Results

### Characteristics of the study population

Of the original 1,426 participants, 1,344 (94.2%) met the criteria for follow-up. Of these, 1,242 (92.4%) were finally contactable via EHRs or telephone interviews. The reasons for exclusion are summarized in Figure 1. The main characteristics of the participants stratified by sex are shown in Table 1. At baseline, the mean age of the study population was 62 years. A high percentage had a family history of DM (31.6%) and a low prevalence of cardiovascular disease (eg, coronary artery disease, stroke, and peripheral artery disease [3.1%, 2%, and 0.8%, respectively]). One-third of the population met the criteria for current smoking, arterial hypertension, and metabolic syndrome, and approximately 50% met the criteria for consumption ≥1 units of alcohol per week. Approximately half of the patients had dyslipidemia, and one in five had regular or poor self-perceived health. The percentage of participants with a high score on the PHQ-9 was 11.6%; this was significantly higher among women. In terms of current treatment, nearly one in four participants were taking statins, and one in five were taking renin-angiotensin system blockers.

**Table 1.** Baseline characteristics of the population studied stratified by sex.

|  | All<br>(N= 1,242) | Men<br>(N= 506) | Women<br>(N=736) | p value |
| --- | --- | --- | --- | --- |
| Age, *mean (SD)* | 61.8 (6.1) | 61.6 (6.3) | 62 (5.9) | 0.214 |
| University studies, n (%) | 395 (31.9) | 222 (44) | 173 (23.5) | **<0.001** |
| Current smoking, n (%) | 438 (35.3) | 238 (47) | 200 (27.2) | **<0.001** |
| Alcohol consumption, n (%) | 598 (48.2) | 349 (69.1) | 249 (33.9) | **<0.001** |
| Family history of DM, n (%) | 392 (31.6) | 148 (29.4) | 244 (33.2) | 0.154 |
| Family history of arterial hypertension, n (%) | 616 (49.8) | 204 (40.5) | 412 (56.3) | **<0.001** |
| Hypertension, n (%) | 400 (32.2) | 185 (36.6) | 215 (29.3) | **0.007** |
| Treated hypertension, n (%) | 360 (90.0) | 169 (91.4) | 191 (88.9) | 0.499 |
| Coronary artery disease, n (%) | 38 (3.1) | 32 (6.3) | 6 (0.8) | **<0.001** |
| Peripheral artery disease, n (%) | 10 (0.8) | 8 (1.6) | 2 (0.3) | **0.011** |
| Stroke, n (%) | 25 (2.0) | 13 (2.6) | 12 (1.6) | 0.248 |
| Chronic atrial fibrillation, n (%) | 30 (2.4) | 17 (3.4) | 13 (1.8) | 0.071 |
| Dyslipidemia, n (%) | 562 (45.3) | 226 (44.8) | 336 (45.7) | 0.754 |
| GFR<sub>e</sub> ≤60 mL/min/1.73 m<sup>2</sup>, n (%) | 14 (1.1) | 10 (2.0) | 4 (0.5) | **0.019** |
| Metabolic syndrome, n (%) | 396 (31.9) | 176 (34.8) | 220 (29.9) | 0.069 |
| Renin-angiotensin system blockers, n (%) | 268 (21.6) | 140 (27.7) | 128 (17.4) | **<0.001** |
| Statins, n (%) | 300 (24.2) | 125 (24.7) | 175 (23.8) | 0.708 |
| Aspirin, n (%) | 76 (6.1) | 50 (9.9) | 26 (3.5) | **0.004** |
| Anticoagulants, n (%) | 24 (1.9) | 13 (2.6) | 11 (1.5) | 0.176 |
| Waist circumference, cm *mean (SD)* | 93.9 (11.9) | 99.9 (9.7) | 89.7 (11.5) | **<0.001** |
| BMI, kg/m<sup>2</sup> *mean (SD)* | 28 (4.5) | 28.4 (3.7) | 27.8 (5) | **0.010** |
| Systolic blood pressure, mmHg, *mean (SD)* | 123.4 (16.8) | 121 (17.3) | 126.8 (15.5) | **<0.001** |
| Diastolic blood pressure, mmHg, *mean (SD)* | 76.8 (9.8) | 78.5 (9.7) | 75.7 (9.7) | **<0.001** |
| Plasma glucose 0 h, mg/dl *mean (SD)* | 100.6 (10.4) | 103.6 (10.5) | 98.5 (9.8) | **<0.001** |
| Plasma glucose 2 h, mg/dl *mean (SD)* | 115.2 (30.6) | 120.3 (31.2) | 111.6 (29.6) | **<0.001** |
| HbA1c, % *mean (SD)* | 5.7 (0.3) | 5.6 (0.4) | 5.7 (0.3) | **0.006** |
| **Psychological status** |  |  |  |  |
| PHQ-9 score, *mean (SD)* | 4.2 (4.4) | 3.2 (4) | 4.9 (4.6) | **<0.001** |
| PHQ-9≥10 (high risk), n (%) | 144 (11.6) | 39 (7.7) | 105 (14.3) | **<0.001** |
| **Health perception (self-reported)** |  |  |  |  |
| Excellent, n (%) | 36 (2.9) | 23 (4.6) | 13 (1.8) | **<0.001** |
| Very good, n (%) | 248 (20.1) | 121 (24.0) | 17.5 (127) |  |
| Good, n (%) | 702 (57) | 287 (56.8) | 415 (57.2) |  |
| Regular, n (%) | 235 (19.1) | 71 (14.1) | 164 (22.6) |  |
| Bad, n (%) | 10 (0.8) | 3 (0.6) | 7 (1.0) |  |
| **Diabetes risk scores** |  |  |  |  |
| FINDRISC, *mean (SD)* | 11.1 (4.3) | 10.8 (4.2) | 11.2 (4.3) | 0.117 |
| DESIR, *mean (SD)* | 11.8 (1.2) | 11.9 (1.2) | 11.8 (1.2) | 0.777 |
| ADA, *mean (SD)* | 5.1 (1.5) | 5.8 (1.3) | 4.6 (1.5) | **<0.001** |

### Incidence of T2DM

During 7.3 years (median) of follow-up, 104 participants (8.4%; 95% CI, 6.8 to 9.9) developed T2DM. Table 2 shows the differences between participants with and without incident T2DM for the main characteristics examined. The risk factors for which values were significantly higher in the group with incident T2DM were hypertension, metabolic syndrome, BMI, waist circumference, systolic and diastolic blood pressure, fasting plasma glucose, OGTT result, HbA1c, impaired glucose tolerance, self-administered PHQ-9 score, and diabetes risk scores (FINDRISC, DESIR, and ADA).

**Table 2.**
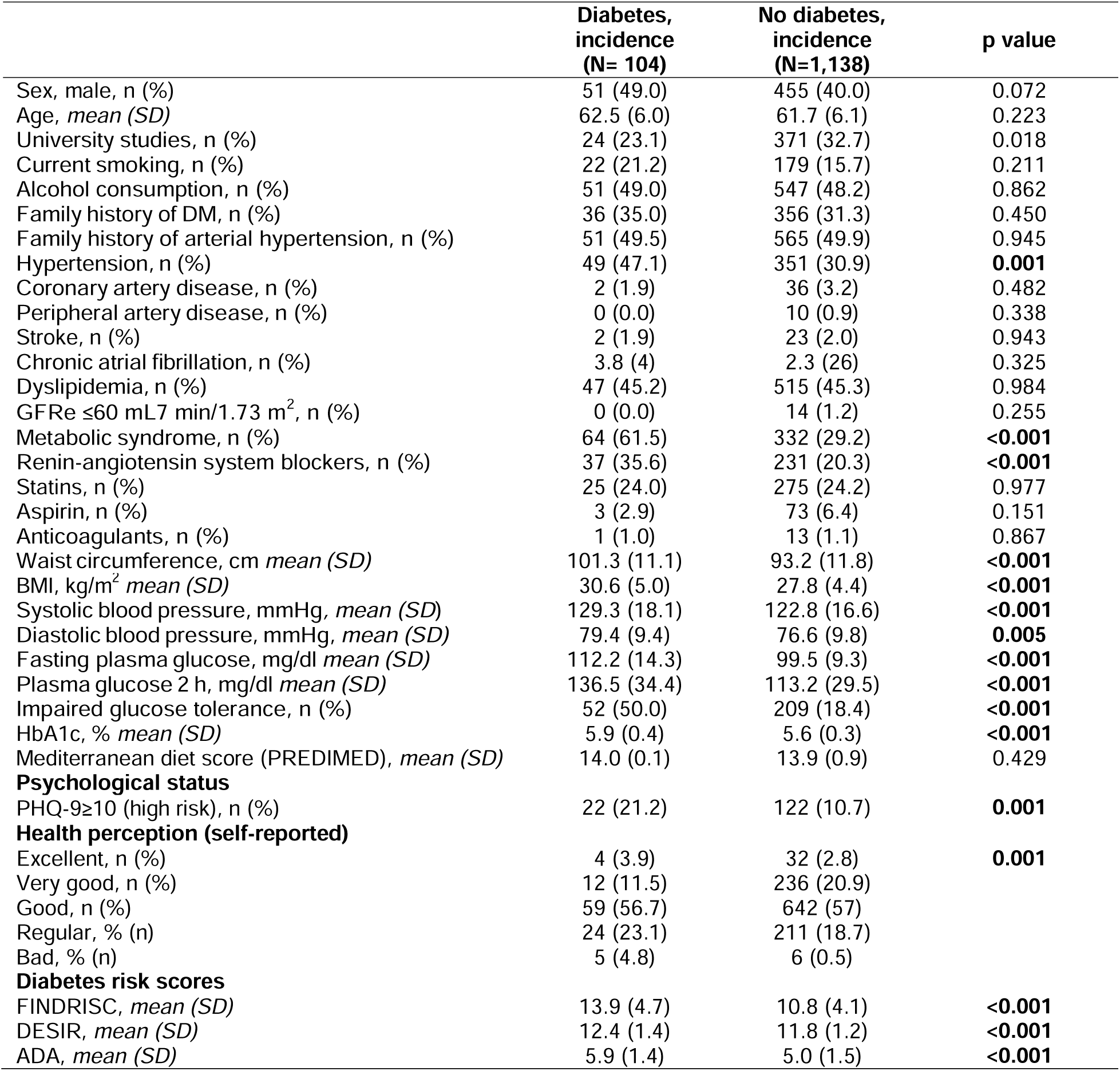
Baseline characteristics of the study population stratified by incidence of diabetes/no diabetes (median 7.3 years of follow-up)

Patients treated with renin-angiotensin system blockers were statistically significantly more likely to be in the diabetes group.

Diabetes risk scores usually include questions about lifestyle, diet, and medical history. Table 3 shows the differences between the two groups. The group without incident T2DM was more likely than the group with incident T2DM to perform at least 30 minutes of physical activity, to eat vegetables, fruit, or berries every day, to have never taken medication for high blood pressure, to have never been diagnosed with high blood sugar, and to have never had gestational diabetes.

**Table 3.**
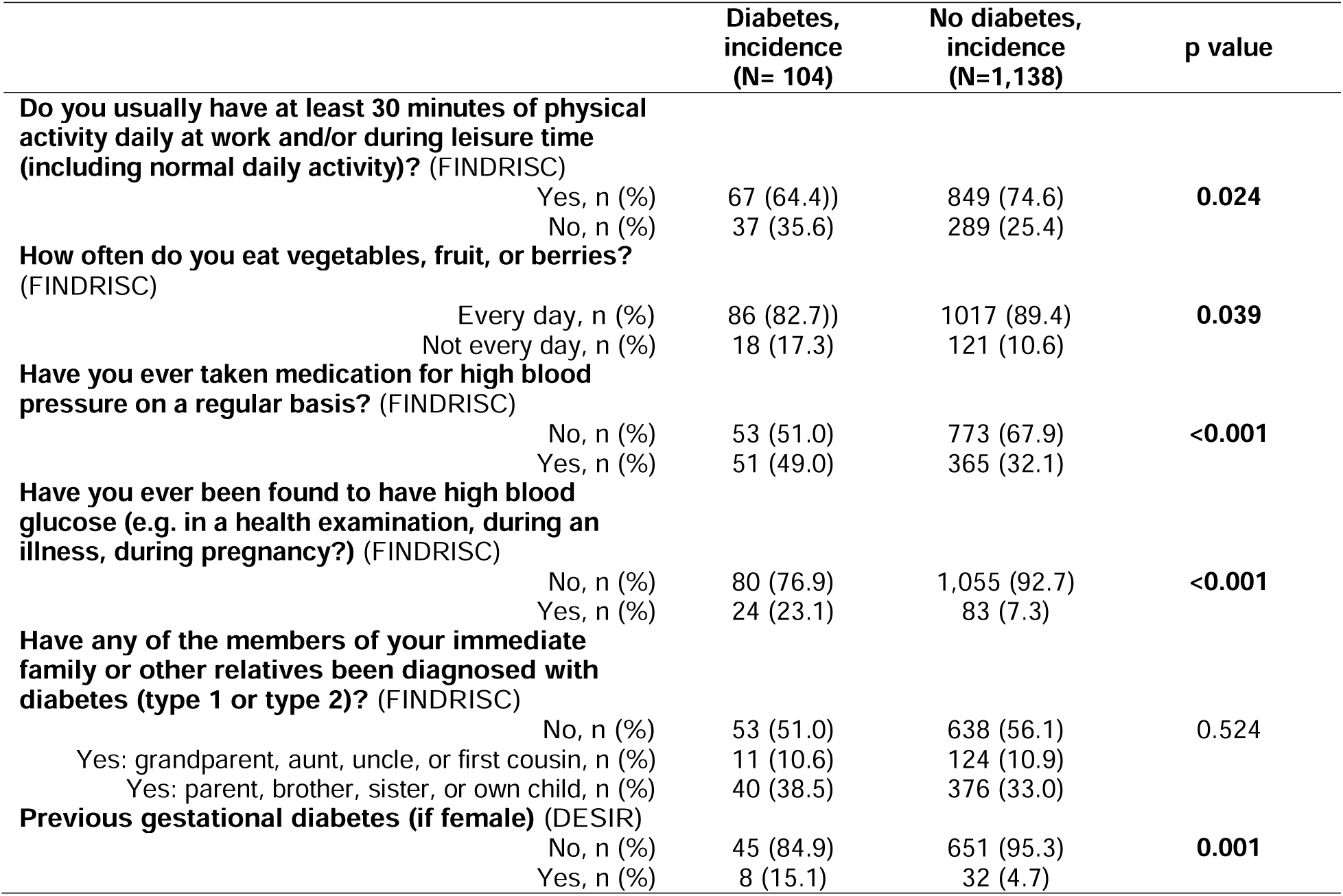
Questions on lifestyle, diet, and medical history included in questionnaires: differences by incidence of diabetes/no diabetes.

|  | **Diabetes,**<br>**incidence**<br>**(N= 104)** | **No diabetes,**<br>**incidence**<br>**(N=1,138)** | **p value** |
| --- | --- | --- | --- |
| **Do you usually have at least 30 minutes of physical activity daily at work and/or during leisure time (including normal daily activity)? (FINDRISC)** |  |  |  |
| Yes, n (%) | 67 (64.4)) | 849 (74.6) | **0.024** |
| No, n (%) | 37 (35.6) | 289 (25.4) |  |
| **How often do you eat vegetables, fruit, or berries? (FINDRISC)** |  |  |  |
| Every day, n (%) | 86 (82.7)) | 1017 (89.4) | **0.039** |
| Not every day, n (%) | 18 (17.3) | 121 (10.6) |  |
| **Have you ever taken medication for high blood pressure on a regular basis? (FINDRISC)** |  |  |  |
| No, n (%) | 53 (51.0) | 773 (67.9) | **<0.001** |
| Yes, n (%) | 51 (49.0) | 365 (32.1) |  |
| **Have you ever been found to have high blood glucose (e.g. in a health examination, during an illness, during pregnancy?) (FINDRISC)** |  |  |  |
| No, n (%) | 80 (76.9) | 1,055 (92.7) | **<0.001** |
| Yes, n (%) | 24 (23.1) | 83 (7.3) |  |
| **Have any of the members of your immediate family or other relatives been diagnosed with diabetes (type 1 or type 2)? (FINDRISC)** |  |  |  |
| No, n (%) | 53 (51.0) | 638 (56.1) | 0.524 |
| Yes: grandparent, aunt, uncle, or first cousin, n (%) | 11 (10.6) | 124 (10.9) |  |
| Yes: parent, brother, sister, or own child, n (%) | 40 (38.5) | 376 (33.0) |  |
| **Previous gestational diabetes (if female) (DESIR)** |  |  |  |
| No, n (%) | 45 (84.9) | 651 (95.3) | **0.001** |
| Yes, n (%) | 8 (15.1) | 32 (4.7) |  |

### Performance of diabetes risk scores

The performance of the FINDRISC score is shown in Table 4. The best cut-off point was >14, achieving a sensitivity of 47.12% (95% CI, 37.2 to 57.2), specificity of 81.37% (95% CI, 79 to 83.6), and positive likelihood ratio of 2.53 (95% CI, 2.0 to 3.21). The AUROC was 0.68 (95% CI, 0.65 to 0.71).

**Table 4.**
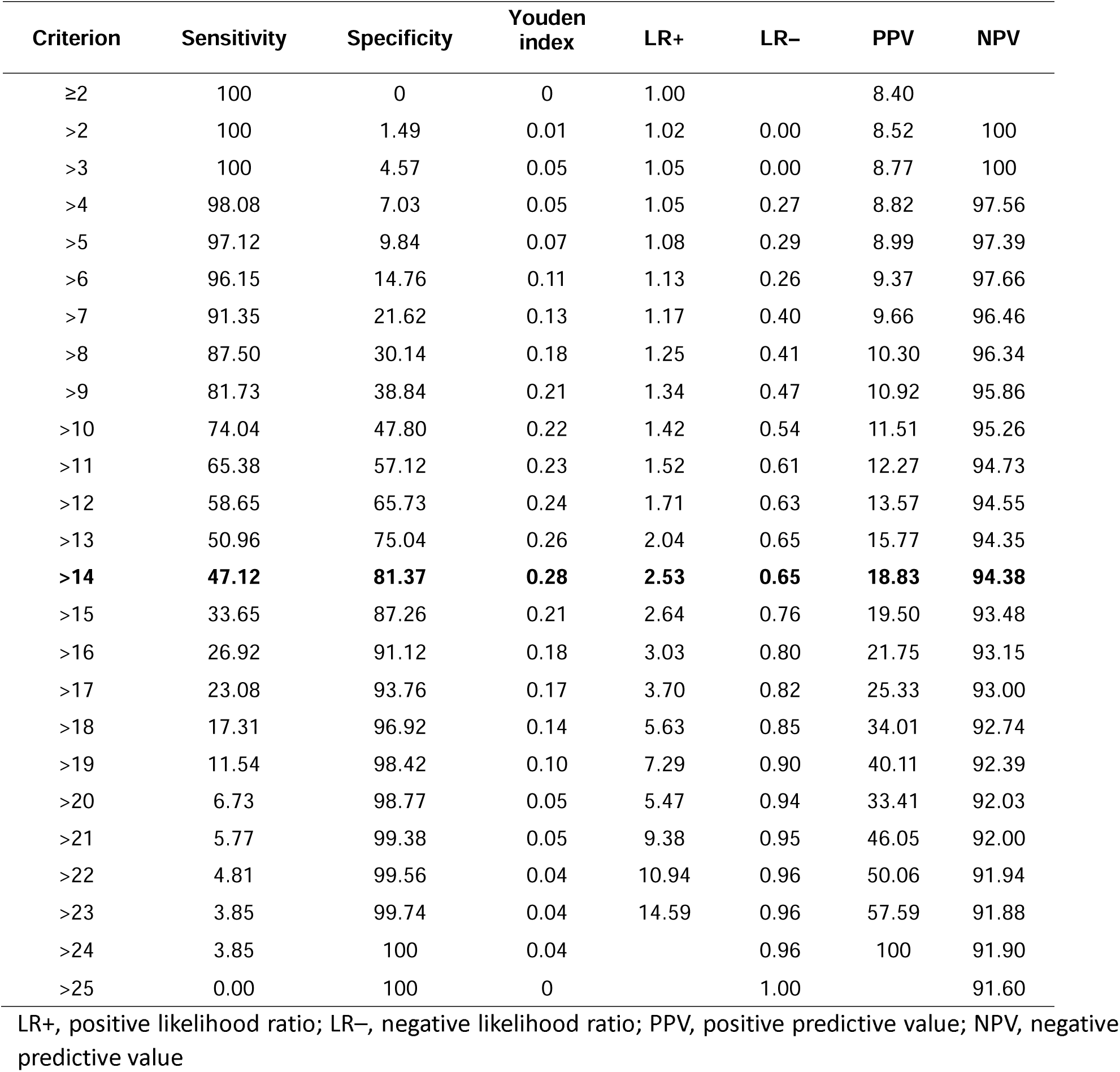
Performance of the FINDRISC diabetes risk score in predicting incident diabetes mellitus after 7.3 years (median) of follow-up.

| Criterion | Sensitivity | Specificity | Youden index | LR+ | LR– | PPV | NPV |
| --- | --- | --- | --- | --- | --- | --- | --- |
| ≥2 | 100 | 0 | 0 | 1.00 |  | 8.40 |  |
| >2 | 100 | 1.49 | 0.01 | 1.02 | 0.00 | 8.52 | 100 |
| >3 | 100 | 4.57 | 0.05 | 1.05 | 0.00 | 8.77 | 100 |
| >4 | 98.08 | 7.03 | 0.05 | 1.05 | 0.27 | 8.82 | 97.56 |
| >5 | 97.12 | 9.84 | 0.07 | 1.08 | 0.29 | 8.99 | 97.39 |
| >6 | 96.15 | 14.76 | 0.11 | 1.13 | 0.26 | 9.37 | 97.66 |
| >7 | 91.35 | 21.62 | 0.13 | 1.17 | 0.40 | 9.66 | 96.46 |
| >8 | 87.50 | 30.14 | 0.18 | 1.25 | 0.41 | 10.30 | 96.34 |
| >9 | 81.73 | 38.84 | 0.21 | 1.34 | 0.47 | 10.92 | 95.86 |
| >10 | 74.04 | 47.80 | 0.22 | 1.42 | 0.54 | 11.51 | 95.26 |
| >11 | 65.38 | 57.12 | 0.23 | 1.52 | 0.61 | 12.27 | 94.73 |
| >12 | 58.65 | 65.73 | 0.24 | 1.71 | 0.63 | 13.57 | 94.55 |
| >13 | 50.96 | 75.04 | 0.26 | 2.04 | 0.65 | 15.77 | 94.35 |
| **>14** | **47.12** | **81.37** | **0.28** | **2.53** | **0.65** | **18.83** | **94.38** |
| >15 | 33.65 | 87.26 | 0.21 | 2.64 | 0.76 | 19.50 | 93.48 |
| >16 | 26.92 | 91.12 | 0.18 | 3.03 | 0.80 | 21.75 | 93.15 |
| >17 | 23.08 | 93.76 | 0.17 | 3.70 | 0.82 | 25.33 | 93.00 |
| >18 | 17.31 | 96.92 | 0.14 | 5.63 | 0.85 | 34.01 | 92.74 |
| >19 | 11.54 | 98.42 | 0.10 | 7.29 | 0.90 | 40.11 | 92.39 |
| >20 | 6.73 | 98.77 | 0.05 | 5.47 | 0.94 | 33.41 | 92.03 |
| >21 | 5.77 | 99.38 | 0.05 | 9.38 | 0.95 | 46.05 | 92.00 |
| >22 | 4.81 | 99.56 | 0.04 | 10.94 | 0.96 | 50.06 | 91.94 |
| >23 | 3.85 | 99.74 | 0.04 | 14.59 | 0.96 | 57.59 | 91.88 |
| >24 | 3.85 | 100 | 0.04 |  | 0.96 | 100 | 91.90 |
| >25 | 0.00 | 100 | 0 |  | 1.00 |  | 91.60 |

LR+, positive likelihood ratio; LR–, negative likelihood ratio; PPV, positive predictive value; NPV, negative predictive value

The FINDRISC-MOOD diabetes risk score, that is, the original FINDRISC plus five points if PHQ-9 >10, showed the same cut-off point as FINDRISC, although the sensitivity and specificity were more balanced (56.7% and 76.7%, respectively) (Table 5). The negative predictive value was 95.08%, slightly higher than that of the original FINDRISC score. The AUROC curve increased to 0.70 (95% CI, 0.67 to 0.72).

**Table 5.**
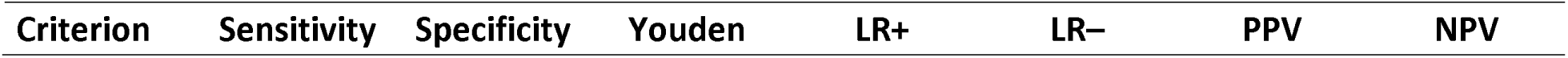
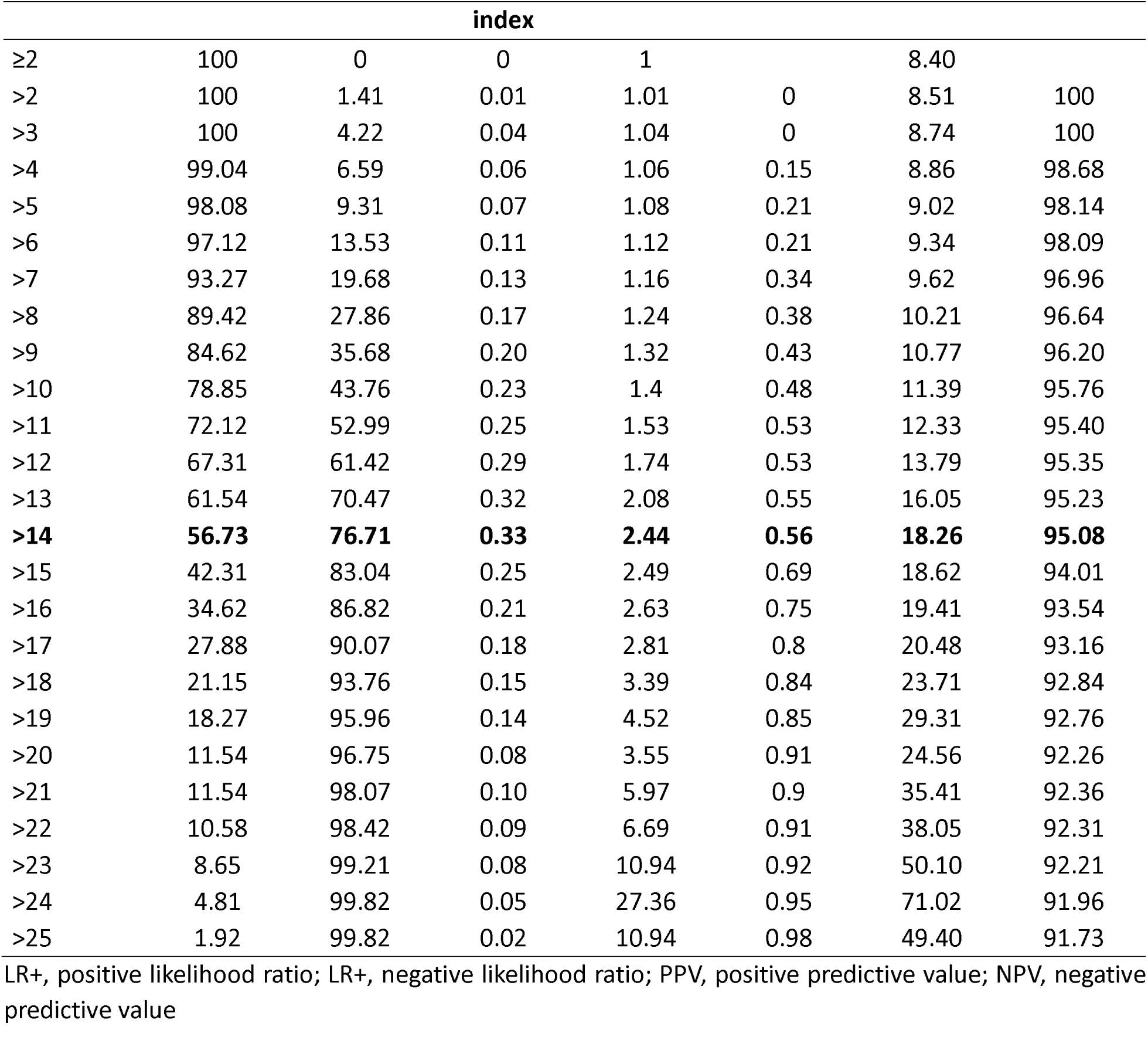
Performance of the FINDRISC-MOOD questionnaire in predicting incident diabetes mellitus after 7.3 years (median) of follow-up.

Finally, the performance of DESIR and ADA is shown in Table 6. The best cut-off points were >12 and >5, respectively. The AUC of the ROC curve was almost equal, with 0.66 (95% CI, 0.63 to 0.68) for DESIR and 0.661 (95% CI, 0.63 to 0.69) for ADA.

**Table 6.** Performance of the DESIR and ADA questionnaires in predicting incident diabetes mellitus after 7.3 years (median) of follow-up.

| DESIR |  |  |  |  |  |  |  |
| --- | --- | --- | --- | --- | --- | --- | --- |
| Criterion | Sensitivity | Specificity | Youden index | LR+ | LR- | PPV | NPV |
| ≥1 | 100 | 0 | 0 | 1.00 |  | 8.40 |  |
| >1 | 100 | 0.088 | 0 | 1.00 | 0.00 | 8.41 | 100 |
| >2 | 99.04 | 0.088 | -0.01 | 0.99 | 10.94 | 8.33 | 49.99 |
| >3 | 99.04 | 0.18 | -0.01 | 0.99 | 5.47 | 8.34 | 67.16 |

| >9 | 98.08 | 2.46 | 0.01 | 1.01 | 0.78 | 8.44 | 93.32 |
| --- | --- | --- | --- | --- | --- | --- | --- |
| >10 | 96.15 | 12.13 | 0.08 | 1.09 | 0.32 | 9.12 | 97.17 |
| >11 | 83.65 | 37.52 | 0.21 | 1.34 | 0.44 | 10.93 | 96.16 |
| **>12** | **51.92** | **71.44** | **0.23** | **1.82** | **0.67** | **14.29** | **94.19** |
| >13 | 13.46 | 95.17 | 0.09 | 2.79 | 0.91 | 20.35 | 92.30 |
| >14 | 0 | 100 | 0 |  | 1.00 |  | 91.60 |
| **ADA** |  |  |  |  |  |  |  |
| Criterion | Sensitivity | Specificity | Youden index | LR+ | LR– | PPV | NPV |
| ≥1 | 100 | 0 | 0 | 1.00 |  | 8.40 |  |
| >1 | 100 | 0.088 | 0 | 1.00 | 0.00 | 8.41 | 100 |
| >2 | 99.04 | 4.75 | 0.04 | 1.04 | 0.20 | 8.71 | 98.18 |
| >3 | 95.19 | 15.11 | 0.10 | 1.12 | 0.32 | 9.32 | 97.16 |
| >4 | 83.65 | 36.12 | 0.20 | 1.31 | 0.45 | 10.72 | 96.01 |
| **>5** | **62.50** | **62.65** | **0.25** | **1.67** | **0.60** | **13.30** | **94.80** |
| >6 | 34.62 | 82.95 | 0.18 | 2.03 | 0.79 | 15.70 | 93.26 |
| >7 | 14.42 | 94.90 | 0.09 | 2.83 | 0.90 | 20.59 | 92.36 |
| >8 | 1.92 | 99.12 | 0.01 | 2.19 | 0.99 | 16.67 | 91.68 |
| >9 | 0 | 100 | 0 |  | 1.00 |  | 91.60 |

LR+, positive likelihood ratio; LR–, negative likelihood ratio; PPV, positive predictive value; NPV, negative predictive value

The AUROCs for each score are also provided in Fig. 2, where a higher value corresponds to the FINDRISC-MOOD risk score, followed by FINDRISC, ADA, and DESIR in order from highest to lowest.

**Figure 2.**
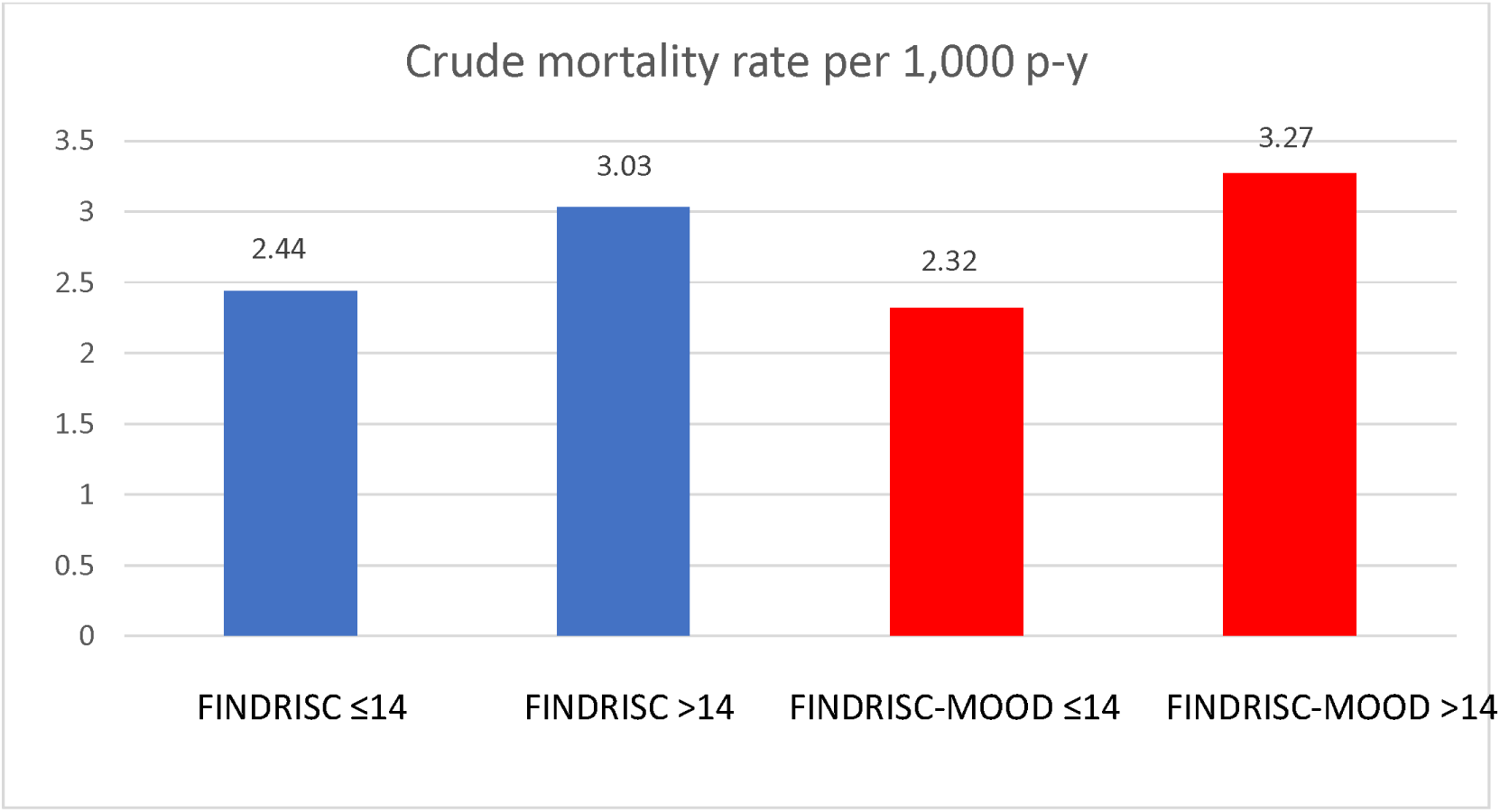
Mortality rates per 1,000 person-years according to FINDRISC and FINDRISC-MOOD.

The results of the bivariate comparisons of the AUROCs are provided in Supplementary Table 1. The differences between values were not statistically significant. The greatest difference was between the FINDRSC-MOOD and DESIR scores (z=1.841, p=0.0657).

### Mortality

There were 24 deaths during follow-up, that is, a crude mortality rate of 2.57 (95% CI, 1.64 to 3.82) per 1,000 person-years. When stratified by FINDRISC score, those with a FINDRISC ≤14 had a crude mortality rate of 2.44 (95% CI, 1.45 to 3.86) per 1,000 person-years and those with a score >14 had a crude mortality rate of 3.03 (95% CI, 1.11 to 6.61) per 1,000 person-years. The rate ratio was 1.24 (95% CI, 0.41 to 3.27), p=0.628.

The crude mortality rate among PHQ-9 negative participants was 2.42 (95% CI, 1.48 to 3.73), and the crude mortality rate among PHQ-9 positive participants was 3.70 (95% CI, 1.01 to 9.48). The rate ratio between the two groups was 1.53 (95% CI, 0.38 to 4.57); p= 0.434.

Similar results were observed with FINDRISC-MOOD. Those with a score below 14 had a crude mortality rate of 2.32 (95% CI, 1.32 to 3.76) per 1,000 person-years, and those with a score >14 had a crude mortality rate of 3.27 (95% CI, 1.41 to 6.45) per 1,000 person-years. The rate ratio was 1.41 (95% CI, 0.52 to 3.50), p=0.427. In this sense, a FINDRISC-MOOD score >14 indicates a slight increase in crude mortality compared to the same score in the traditional FINDRISC questionnaire, probably due to the increased mortality risk in PHQ-9 positive individuals, as we found (Figure 2).

## Discussion

Of the 1,242 participants in the cohort study, 104 (8.4%; 95% CI, 6.8 to 9.9) developed T2DM during the follow-up period. The ability of the questionnaires to detect incident diabetes in the long term, according to the established cut-off point, was assessed based on the sensitivity. In this study, sensitivity ranged from 47.12% (95% CI, 37.2 to 57.2) for the original FINDRISC ≥ 14 to 62.50% (95%, 52.5 to 71.8) for ADA > 5.

The FINDRISC-MOOD questionnaire was more sensitive than the original FINDRISC and would therefore be preferred for screening. When dealing with a treatable disease where it is desirable to diagnose before complications develop, as in the case of diabetes, it is preferable to choose cut-off points with high sensitivity.

In this sense, effective screening would require a sensitivity of at least 80%. Therefore, the best cut-off for this purpose would vary in the FINDRISC-related questionnaires: ≥10 for FINDRISC-MOOD and ≥9 for the original FINDRISC. This approach could prove inconvenient, because high sensitivity leads to worse specificity and, therefore, a higher percentage of false positives (1-specificity), which, in addition to the possible psychological damage caused to patients by an incorrect diagnosis, could have led to unnecessary pharmacological treatment, such as metformin (24) and semaglutide (25). For this reason, it is always necessary for laboratory measurements to subsequently detect possible false positives. The worst specificity was for ADA > 5, with 62.65% (95% CI, 59.8 to 65.5), and the highest specificity was for FINDRISC ≥ 14, with 81.37% (95% CI, 79 to 83.6).

Although FINDRISC-MOOD showed a greater discriminative capacity (AUROC) than the other questionnaires, the differences were not significant. Several authors have improved the AUROC of predictive risk scores by adding laboratory measurements such as fasting plasma glucose (26) (27), HbA1c alone (28), and HbA1c plus fasting plasma glucose (29). This strategy improves discriminatory accuracy but hampers the objective of a quick and noninvasive prediction using easily measured variables. The predictive diabetes risk scores could be considered a prescreening tool to identify which screened individuals could benefit from measurement of fasting plasma glucose or HbA1c. This approach constitutes a relatively cost-effective screening program, as highlighted by the International Diabetes Federation (30).

Owing to differences in lifestyle and the prevalence of chronic diseases such as obesity, prediabetes, and diabetes between communities and ethnicities, it is often necessary to calculate the optimal cut-off point for diabetes risk scores for each country. From a statistical point of view, the best cut-off point is the one that achieves a higher Youden index, although as previously mentioned, this strategy can be modified depending on the purpose of screening. According to the highest Youden index, the best cut-off points for the FINDRISC and FINDRISC-MOOD questionnaires are the same (≥14).

The cut-offs used change over time and according to study site. In Europe, the original FINDRISC designed by Linström and Tuomilheto in Finland (9) showed an optimal cut-off ≥9 in two consecutive cohorts after 10 years of follow-up with the following performance parameters (Youden index, sensitivity, specificity, and AUC) 0.59, 78%, 81%, and 0.85 in the 1987 cohort and 0.53, 77%, 76%, and 0.87 in the 1992 cohort. In the Norwegian general population aged over 20 years, the best FINDRISC cut-off, according to the highest Youden index, was ≥11 (sensitivity, 73%; specificity, 67%) after 10 years of follow-up. The AUC-ROC was 0.77 (31).

The study by Alssema et al. (2008) with subjects from the HOORN (32) (33) (n= 1,434), PREVEND (34) (35) (n= 2,713), and MORGEN (36) (37) (n= 863) cohorts in the Netherlands included six variables from FINDRISC (age, BMI, waist circumference, use of blood pressure medication, history of high blood glucose, and family history of diabetes mellitus). The total score of the orginal FINDRISC ranged from 0 to 22, and it yielded two acceptable cut-offs for each cohort. Values (Youden index, sensitivity, and specificity) were as follows: HOORN cohort, 0.28, 52%, and 76% for a cut-off ≥10 and 0.26, 84%, and 42% for a cut-off ≥7; PREVEND cohort, 0.28, 43%, and 85% for a cut-off ≥10 and 0.42, 78%, and 64% for a cut-off ≥7; MORGEN cohort, 0.30, 47%, and 83% for a cut-off ≥10 and 0.27, 75%, and 52% for a cut-off ≥7 (38). The second study by Alssema et al. (2011) included the previous variables used in 2008, as well as sex and smoking, with an optimal cut-off of ≥7 (Youden index, 0.39; sensitivity, 76%; specificity, 63%; AUROC, 0.74) (39).

A simplified version of the FINDRISC without the diet and physical activity variables in the questionnaire was tested in Germany over three years. The total score of the modified FINDRISC ranged from 1 to 23. The best cut-off was ≥9 (Youden index, 0.406; sensitivity, 73.5%; specificity, 67.1%; AUROC, 0.77) (40).

In Spain, a population-based prospective study performed in the town of Pizarra (Málaga) followed 824 individuals for six years to evaluate the performance of FINDRISC. All participants without known diabetes underwent an OGTT at baseline and at the end of follow-up. The best prediction of the risk of incident T2DM was found in subjects with a FINDRISC cut-off of nine (OR, 19.37; 95% CI, 8.86 to 42.34; P <0.0001) and an AUROC of 0.75. No information was provided on sensitivity, specificity, or the Youden index (41).

In Asia, an Iranian cohort study of 1,537 first-degree relatives of consecutive individuals with T2DM aged 30-70 years with 7.5 years of follow-up showed the best FINDRISC cut-off to be ≥13 (Youden index, 0.375; sensitivity, 83.6%; specificity, 53.9%) (42). In a rural adult Chinese population, FINDRISC showed a surprisingly low cut-off of ≥5 (Youden index, 0.254; sensitivity, 54.3%; specificity, 71.1%) for predicting the incidence of T2DM after one year of follow-up (43).

In contrast to most studies, the AUC-ROC of the Pizarra study was closer to our results, probably owing to a common lifestyle pattern and a progressive decrease in discriminative ability, as is relatively common in contemporary studies. The comparison of AUROCs showed p values less than 0.10 between FINDRISC-MOOD and DESIR and ADA and very close to 0.05 between FINDRISC-MOOD and ADA. These results point to FINDRISC-MODD as being more useful than the other two questionnaires.

The addition of five points to the original FINDRISC when the PHQ-9 questionnaire score is greater than 10 points (44) led to an improvement in AUROC, which, although not significantly better than the original FINDRISC score, provides two essential advantages: first, it reaches a value of 0.70, the minimum to consider that a test provides sufficient discrimination; and second, a notable improvement with respect to DESIR and ADA, which does not reach statistical significance but is close to it. Furthermore, the PHQ-9 questionnaire can be used with the FINDRISC questionnaire because it is self-administered and requires relatively little time. This is an exciting contribution because depression should be screened for regularly, as recommended by the US Preventive Services Task Force (45). In addition, the ADA recommends screening for depression in patients with diabetes (46). Furthermore, depression is associated with up to a 65% increased risk of DM (4) (47), making addition of the PHQ-9 particularly interesting when assessing risk of DM. Furthermore, the PHQ-9 can be considered a first-line tool for the diagnosis of depression in primary care settings owing to its ease of administration, good acceptability, and sensitivity for detecting depression (45).

Our study has several limitations. First, the incidence of DM was not measured using the same method as at baseline (OGTT). However, as we used four sources of data (self-reported diagnosis, diagnosis by a general practitioner, baseline plasma glucose levels, and use of hypoglycemic medication) that were consistent with other authors (48) (49), information bias is unlikely. Second, the FINDRISC score estimates the risk of participants aged 35 to 64 years developing T2DM within 10 years; our study included some patients aged 65 years or older with a follow-up period of 7.3 years. This may have altered the accuracy of the results. Third, the diabetes risk scores in our study were performed on a representative population in the north of the city of Madrid. However, lifestyle, diet, and prevalence of obesity may differ from the rest of the city, given the lower gross domestic product in the southern area. This may limit extrapolation to these areas or other Spanish cities.

Our study also has important strengths. The study population reflects the average risk of DM, without a large number of people at high risk of developing T2DM, as in other studies carried out in hospital samples (28) (50), patients with chronic infectious diseases (51), and regions with a high prevalence of sedentary lifestyle and obesity (28). Another strength is that the extension of the original FINDRISC, FINDRISC-MOOD, is an easily administered tool that does not require an extra investment in health professionals, because the PHQ-9 is typically self-administered (52).

Conclusion: Of the four diabetes risk scores used, FINDRISC-MOOD performed best. The differences between the scores were small and not statistically significant. However, the use of FINDRISC-MOOD adds value to the assessment of diabetes risk in patients with high risk of depression as assessed by the PHQ-9 questionnaire. This dual screening strategy is advantageous because individuals are prognostically stratified according to two common conditions: depression and diabetes, both of which are associated with all-cause mortality with crude mortality rates ranging from 3.03 to 3.70 per 1,000 person-years. FINDRISC-MOOD could be used to identify the risk of future T2DM in primary care.

## Data availability statement

Data are available upon reasonable request.

## Ethics statements

### Patient consent for publication

Not required

### Ethics approval

The SPREDIA-2 study was approved by the Research Ethics Committee (EC) of Hospital Carlos III (Madrid) in 2010. Authorization to review the clinical data of SPREDIA-2 study participants from their primary care electronic health record (PC-EHR) and to conduct a telephone interview during the last year of follow-up was obtained from the La Paz Hospital EC in 2017. Participants gave informed consent to participate in the study prior to enrollment.

## Data Availability

Data are available upon reasonable request

## Acknowledgements

We would like to thank all members of the SPREDIA-2 Group: Fernando Laguna (Hospital Carlos III), Pedro Fernández-García (Hospital Carlos III), Luis Montesano-Sánchez (Hospital Carlos III), Pedro Patrón (Hospital Carlos III), Leopoldo Pérez-Isla (Hospital Clínico de San Carlos), David Vicent (Hospital Carlos III), Ignacio Vicente (CS Monóvar), Sara Artola (CS M^a^ Jesús Hereza), M^a^ Isabel Granados-Menéndez (CS Monóvar), Domingo Beamud-Victoria (CS Felipe II), Isidoro Dujovne-Kohan (CS Los Castillos), Rosa María Chico-Moraleja (Hospital Central de la Defensa), Carmen Martín-Madrazo (CS Monóvar), Rosario Echegoyen de Nicolás (CS Benita de Ávila), Concepción Aguilera Linde (CS Ciudad Periodistas), Álvaro R Aguirre De Carcer Escolano (CS La Ventilla), Patricio Alonso Sacristán (CS Ciudad Periodistas), M Jesús Álvarez Otero (CS Dr Castroviejo), Paloma Arribas Pérez (CS Santa Hortensia), Maria Luisa Asensio Ruiz (CS Fuentelarreina), Pablo Astorga Díaz (CS Barrio Pilar), Begoña Berriatua Ena (CS Dr Castroviejo), Ana Isabel Bezos Varela (CS José Marva), María José Calatrava Triguero (CS Ciudad Jardín), Carlos Casanova García (CS Barrio Pilar), Ángeles Conde Llorente (CS Barrio Pilar), Concepción Díaz Laso (CS Fuentelarreina), Emilia Elviro García (CS Ciudad Periodistas), Orlando Enríquez Dueñas (CS Fuentelarreina), María Isabel Ferrer Zapata (CS El Greco), Froilán Antuña (CS Ciudad Periodistas), Maria Isabel García Lazaro (CS Ciudad Periodistas), Maria Teresa Gómez Rodríguez (CS Barrio Pilar), África Gómez Lucena (CS La Ventilla), Francisco Herrero Hernández (CS La Ventilla), Rosa Julián Viñals (CS Dr Castroviejo), Gerardo López Ruiz Ogarrio “in memoriam” (CS Barrio Pilar), Maria Del Carmen Lumbreras Manzano (CS José Marva), Sonsoles Paloma Luquero López (CS Ciudad Periodistas), Ana Martínez Cabrera Peláez (CS Barrio Pilar), Montserrat Nieto Candenas (CS La Ventilla), María Alejandra Rabanal Carrera (CS Barrio Pilar), Ángel Castellanos Rodríguez (CS Ciudad Periodistas), Ana López Castellanos (CS La Ventilla), Milagros Velázquez García (CS Barrio Pilar) and Margarita Ruiz Pacheco (CS Dr. Castroviejo).

## Author Contributors

MASF, CBL, PGC, FJSR: conceptualization, methodology, validation, formal analysis, writing—original draft, visualisation.

JM, CL, BFP, EEC, FGI, TGA: investigation and resources

VCR, VSA, CSR, SLL: supervision and validation

VIC, BTE: review and editing

JCV: data curation and supervision

FRA: validation, final review and editing

## References

1. Yu H jie, Ho M, Liu X, Yang J, Chau PH, Fong DYT. Association of weight status and the risks of diabetes in adults: a systematic review and meta-analysis of prospective cohort studies. Int J Obes. 2022 Jun;46(6):1101–13.

2. DeFronzo RA, Ferrannini E, Groop L, Henry RR, Herman WH, Holst JJ, et al. Type 2 diabetes mellitus. Nat Rev Dis Primer. 2015 Jul 23;1:15019.

3. Luo H, Huang Y, Zhang Q, Yu K, Xie Y, Meng X, et al. Impacts of physical activity and particulate air pollution on the onset, progression and mortality for the comorbidity of type 2 diabetes and mood disorders. Sci Total Environ. 2023 Sep 10;890:164315.

4. Mezuk B, Eaton WW, Albrecht S, Golden SH. Depression and type 2 diabetes over the lifespan: a meta-analysis. Diabetes Care. 2008 Dec;31(12):2383–90.

5. Emerging Risk Factors Collaboration, Sarwar N, Gao P, Seshasai SRK, Gobin R, Kaptoge S, et al. Diabetes mellitus, fasting blood glucose concentration, and risk of vascular disease: a collaborative meta-analysis of 102 prospective studies. Lancet Lond Engl. 2010 Jun 26;375(9733):2215–22.

6. Alicic RZ, Rooney MT, Tuttle KR. Diabetic Kidney Disease: Challenges, Progress, and Possibilities. Clin J Am Soc Nephrol CJASN. 2017 Dec 7;12(12):2032–45.

7. Raghavan S, Vassy JL, Ho Y, Song RJ, Gagnon DR, Cho K, et al. Diabetes Mellitus–Related All-Cause and Cardiovascular Mortality in a National Cohort of Adults. J Am Heart Assoc. 2019 Feb 19;8(4):e011295.

8. Son JW, Jang EH, Kim MK, Kim IT, Roh YJ, Baek KH, et al. Diabetic retinopathy is associated with subclinical atherosclerosis in newly diagnosed type 2 diabetes mellitus. Diabetes Res Clin Pract. 2011 Feb;91(2):253–9.

9. Lindström J, Tuomilehto J. The Diabetes Risk Score. Diabetes Care. 2003 Mar 1;26(3):725–31.

10. Balkau B, Lange C, Fezeu L, Tichet J, De Lauzon-Guillain B, Czernichow S, et al. Predicting Diabetes: Clinical, Biological, and Genetic Approaches. Diabetes Care. 2008 Oct 1;31(10):2056–61.

11. American Diabetes Association. 2. Classification and Diagnosis of Diabetes: Standards of Medical Care in Diabetes-2020. Diabetes Care. 2020 Jan;43(Suppl 1):S14–31.

12. Vancampfort D, Mitchell AJ, De Hert M, Sienaert P, Probst M, Buys R, et al. TYPE 2 DIABETES IN PATIENTS WITH MAJOR DEPRESSIVE DISORDER: A META-ANALYSIS OF PREVALENCE ESTIMATES AND PREDICTORS. Depress Anxiety. 2015 Oct;32(10):763–73.

13. Salinero-Fort MÁ, de Burgos-Lunar C, Mostaza Prieto J, Lahoz Rallo C, Abánades-Herranz JC, Gómez-Campelo P, et al. Validating prediction scales of type 2 diabetes mellitus in Spain: the SPREDIA-2 population-based prospective cohort study protocol. BMJ Open. 2015 Jul 28;5(7):e007195.

14. Spitzer RL. Validation and Utility of a Self-report Version of PRIME-MDThe PHQ Primary Care Study. JAMA. 1999 Nov 10;282(18):1737.

15. Martínez-González MA, García-Arellano A, Toledo E, Salas-Salvadó J, Buil-Cosiales P, Corella D, et al. A 14-Item Mediterranean Diet Assessment Tool and Obesity Indexes among High-Risk Subjects: The PREDIMED Trial. Peiró C, editor. PLoS ONE. 2012 Aug 14;7(8):e43134.

16. Ware J, Kosinski M, Keller SD. A 12-Item Short-Form Health Survey: construction of scales and preliminary tests of reliability and validity. Med Care. 1996 Mar;34(3):220–33.

17. de Burgos-Lunar C, Salinero-Fort MA, Cárdenas-Valladolid J, Soto-Díaz S, Fuentes-Rodríguez CY, Abánades-Herranz JC, et al. Validation of diabetes mellitus and hypertension diagnosis in computerized medical records in primary health care. BMC Med Res Methodol. 2011 Oct 28;11:146.

18. Gil Montalbán E, Ortiz Marrón H, López-Gay Lucio-Villegas D, Zorrilla Torrás B, Arrieta Blanco F, Nogales Aguado P. [Validity and concordance of electronic health records in primary care (AP-Madrid) for surveillance of diabetes mellitus. PREDIMERC study]. Gac Sanit. 2014;28(5):393–6.

19. Riise HKR, Graue M, Igland J, Birkeland KI, Kolltveit BCH. Prevalence of increased risk of type 2 diabetes in general practice: a cross-sectional study in Norway. BMC Prim Care. 2023 Jul 20;24(1):151.

20. Martin A, Rief W, Klaiberg A, Braehler E. Validity of the Brief Patient Health Questionnaire Mood Scale (PHQ-9) in the general population. Gen Hosp Psychiatry. 2006 Jan;28(1):71–7.

21. Kroenke K, Spitzer RL, Williams JBW, Löwe B. The Patient Health Questionnaire Somatic, Anxiety, and Depressive Symptom Scales: a systematic review. Gen Hosp Psychiatry. 2010;32(4):345–59.

22. Costantini L, Pasquarella C, Odone A, Colucci ME, Costanza A, Serafini G, et al. Screening for depression in primary care with Patient Health Questionnaire-9 (PHQ-9): A systematic review. J Affect Disord. 2021 Jan 15;279:473–83.

23. Ford ES, Giles WH, Dietz WH. Prevalence of the metabolic syndrome among US adults: findings from the third National Health and Nutrition Examination Survey. JAMA. 2002 Jan 16;287(3):356–9.

24. Madsen KS, Chi Y, Metzendorf MI, Richter B, Hemmingsen B. Metformin for prevention or delay of type 2 diabetes mellitus and its associated complications in persons at increased risk for the development of type 2 diabetes mellitus. Cochrane Database Syst Rev. 2019 Dec 3;12(12):CD008558.

25. Rubino D, Abrahamsson N, Davies M, Hesse D, Greenway FL, Jensen C, et al. Effect of Continued Weekly Subcutaneous Semaglutide vs Placebo on Weight Loss Maintenance in Adults With Overweight or Obesity: The STEP 4 Randomized Clinical Trial. JAMA. 2021 Apr 13;325(14):1414–25.

26. Bozorgmanesh M, Hadaegh F, Ghaffari S, Harati H, Azizi F. A simple risk score effectively predicted type 2 diabetes in Iranian adult population: population-based cohort study. Eur J Public Health. 2011 Oct;21(5):554–9.

27. Guasch-Ferré M, Bulló M, Costa B, Martínez-Gonzalez MÁ, Ibarrola-Jurado N, Estruch R, et al. A risk score to predict type 2 diabetes mellitus in an elderly Spanish Mediterranean population at high cardiovascular risk. PloS One. 2012;7(3):e33437.

28. Meijnikman AS, De Block CEM, Verrijken A, Mertens I, Van Gaal LF. Predicting type 2 diabetes mellitus: a comparison between the FINDRISC score and the metabolic syndrome. Diabetol Metab Syndr. 2018;10:12.

29. Hippisley-Cox J, Coupland C. Development and validation of QDiabetes-2018 risk prediction algorithm to estimate future risk of type 2 diabetes: cohort study. BMJ. 2017 Nov 20;359:j5019.

30. International Diabetes Federation Guideline Development Group. Global guideline for type 2 diabetes. Diabetes Res Clin Pract. 2014 Apr;104(1):1–52.

31. Jølle A, Midthjell K, Holmen J, Carlsen SM, Tuomilehto J, Bjørngaard JH, et al. Validity of the FINDRISC as a prediction tool for diabetes in a contemporary Norwegian population: a 10-year follow-up of the HUNT study. BMJ Open Diabetes Res Care. 2019;7(1):e000769.

32. Mooy JM, Grootenhuis PA, de Vries H, Valkenburg HA, Bouter LM, Kostense PJ, et al. Prevalence and determinants of glucose intolerance in a Dutch caucasian population. The Hoorn Study. Diabetes Care. 1995 Sep;18(9):1270–3.

33. de Vegt F, Dekker JM, Jager A, Hienkens E, Kostense PJ, Stehouwer CD, et al. Relation of impaired fasting and postload glucose with incident type 2 diabetes in a Dutch population: The Hoorn Study. JAMA. 2001 Apr 25;285(16):2109–13.

34. Pinto-Sietsma SJ, Janssen WMT, Hillege HL, Navis G, Zeeuw DD, Jong PED. Urinary albumin excretion is associated with renal functional abnormalities in a nondiabetic population. J Am Soc Nephrol JASN. 2000 Oct;11(10):1882–8.

35. Verhave JC, Hillege HL, Burgerhof JGM, Gansevoort RT, de Zeeuw D, de Jong PE, et al. The association between atherosclerotic risk factors and renal function in the general population. Kidney Int. 2005 May;67(5):1967–73.

36. Mensink M, Corpeleijn E, Feskens EJM, Kruijshoop M, Saris WHM, de Bruin TWA, et al. Study on lifestyle-intervention and impaired glucose tolerance Maastricht (SLIM): design and screening results. Diabetes Res Clin Pract. 2003 Jul;61(1):49–58.

37. van Dam RM, Boer JM, Feskens EJ, Seidell JC. Parental history of diabetes modifies the association between abdominal adiposity and hyperglycemia. Diabetes Care. 2001 Aug;24(8):1454–9.

38. Alssema M, Feskens EJM, Bakker SJL, Gansevoort RT, Boer JMA, Heine RJ, et al. [Finnish questionnaire reasonably good predictor of the incidence of diabetes in The Netherlands]. Ned Tijdschr Geneeskd. 2008 Nov 1;152(44):2418–24.

39. Alssema M, Vistisen D, Heymans MW, Nijpels G, Glümer C, Zimmet PZ, et al. The Evaluation of Screening and Early Detection Strategies for Type 2 Diabetes and Impaired Glucose Tolerance (DETECT-2) update of the Finnish diabetes risk score for prediction of incident type 2 diabetes. Diabetologia. 2011 May;54(5):1004–12.

40. Bergmann A, Li J, Wang L, Schulze J, Bornstein SR, Schwarz PEH. A simplified Finnish diabetes risk score to predict type 2 diabetes risk and disease evolution in a German population. Horm Metab Res Horm Stoffwechselforschung Horm Metab. 2007 Sep;39(9):677–82.

41. Soriguer F, Valdés S, Tapia MJ, Esteva I, Ruiz de Adana MS, Almaraz MC, et al. [Validation of the FINDRISC (FINnish Diabetes RIsk SCore) for prediction of the risk of type 2 diabetes in a population of southern Spain. Pizarra Study]. Med Clin (Barc). 2012 Apr 14;138(9):371–6.

42. Janghorbani M, Adineh H, Amini M. Finnish Diabetes Risk Score to predict type 2 diabetes in the Isfahan diabetes prevention study. Diabetes Res Clin Pract. 2013 Dec;102(3):202–9.

43. Zhang M, Zhang H, Wang C, Ren Y, Wang B, Zhang L, et al. Development and Validation of a Risk-Score Model for Type 2 Diabetes: A Cohort Study of a Rural Adult Chinese Population. PloS One. 2016;11(4):e0152054.

44. Levis B, Benedetti A, Thombs BD, DEPRESsion Screening Data (DEPRESSD) Collaboration. Accuracy of Patient Health Questionnaire-9 (PHQ-9) for screening to detect major depression: individual participant data meta-analysis. BMJ. 2019 Apr 9;365:l1476.

45. Siu AL, US Preventive Services Task Force (USPSTF), Bibbins-Domingo K, Grossman DC, Baumann LC, Davidson KW, et al. Screening for Depression in Adults: US Preventive Services Task Force Recommendation Statement. JAMA. 2016 Jan 26;315(4):380–7.

46. American Diabetes Association. 4. Lifestyle Management. Diabetes Care. 2017 Jan;40(Suppl 1):S33–43.

47. Campayo A, de Jonge P, Roy JF, Saz P, de la Cámara C, Quintanilla MA, et al. Depressive disorder and incident diabetes mellitus: the effect of characteristics of depression. Am J Psychiatry. 2010 May;167(5):580–8.

48. NCD Risk Factor Collaboration (NCD-RisC). Effects of diabetes definition on global surveillance of diabetes prevalence and diagnosis: a pooled analysis of 96 population-based studies with 331,288 participants. Lancet Diabetes Endocrinol. 2015 Aug;3(8):624–37.

49. Kazemian P, Shebl FM, McCann N, Walensky RP, Wexler DJ. Evaluation of the Cascade of Diabetes Care in the United States, 2005-2016. JAMAn Intern Med. 2019 Oct 1;179(10):1376–85.

50. Jiménez-Lucena R, Rangel-Zúñiga OA, Alcalá-Díaz JF, López-Moreno J, Roncero-Ramos I, Molina-Abril H, et al. Circulating miRNAs as Predictive Biomarkers of Type 2 Diabetes Mellitus Development in Coronary Heart Disease Patients from the CORDIOPREV Study. Mol Ther Nucleic Acids. 2018 Sep 7;12:146–57.

51. Galaviz KI, Schneider MF, Tien PC, Althoff KN, Ali MK, Ofotokun I, et al. Expanding the Finnish Diabetes Risk Score for Predicting Diabetes Incidence in People Living with HIV. AIDS Res Hum Retroviruses. 2021 May;37(5):373–9.

52. Kroenke K, Spitzer RL, Williams JBW. The Patient Health Questionnaire-2: validity of a two-item depression screener. Med Care. 2003 Nov;41(11):1284–92.

