## Supplementary material for "Validation of Diabetes Prediction Scores: Does adding a high risk for depression increase the area under the curve?"

| **FINDRISC-MOOD VS. FINDRISC** | |
| --- | --- |
| **Difference between areas** | 0.0175 |
| **Standard error** | 0.0120 |
| **95% Confidence interval** | -0.00599 to 0.0410 |
| **Z statistic** | 1.460 |
| **Significance level** | P=0.1443 |
| **FINDRISC-MOOD VS. ADA** | |
| **Difference between areas** | 0.0374 |
| **Standard error** | 0.0214 |
| **95% Confidence interval** | -0.00459 to 0.0794 |
| **Z statistic** | 1.746 |
| **Significance level** | P=0.0808 |
| **FINDRISC-MOOD VS. DESIR** | |
| **Difference between areas** | 0.0413 |
| **Standard error** | 0.0224 |
| **95% Confidence interval** | -0.00268 to 0.0852 |
| **Z statistic** | 1.841 |
| **Significance level** | P=0.0657 |
| **FINDRISC VS. ADA** | |
| **Difference between areas** | 0.0199 |
| **Standard error** | 0.0198 |
| **95% Confidence interval** | -0.0190 to 0.0588 |
| **Z statistic** | 1.004 |
| **Significance level** | P=0.3153 |
| **FINDRISC VS. DESIR** | |
| **Difference between areas** | 0.0238 |
| **Standard error** | 0.0223 |
| **95% Confidence interval** | -0.0199 to 0.0674 |
| **Z statistic** | 1.067 |
| **Significance level** | P=0.2860 |
| **ADA VS. DESIR** | |
| **Difference between areas** | 0.00386 |
| **Standard error** | 0.0223 |
| **95% Confidence interval** | -0.0398 to 0.0475 |
| **Z statistic** | 0.173 |
| **Significance level** | P=0.8625 |

**Supplementary Table 1. Pairwise comparison of ROC curves**
